# Assessment of authors understanding of the STROBE-nut reporting guidelines

**DOI:** 10.1101/2020.10.15.20211268

**Authors:** Dana Hawwash, Michelle Seck, Elisa Quaranta, Patrick Kolsteren, Carl Lachat

**Author notes:** Corresponding author: Carl Lachat, Department of Food Technology, Safety and Health, Faculty of Bioscience Engineering, Coupure links 653, 9000 Ghent-Belgium. Declarations *Data Sharing Statement* Annex 1: Data collection template. Annex 2: Invitation letter, Informed consent and information sheet, baseline questionnaires. Annex 3: Skype questionnaire. Annex 4: Examples of Items on the STROBE nut list that was difficult to understand. Annex 5: Anonymised participant-level responses. Competing Interests Authors (CL DH PK,) have developed STROBE-nut for nutritional epidemiology cited in this manuscript. Author Contributions Conceptualization: CL DH PK. Supervision: CL PK. Wrote the first draft of the manuscript: DH. Contributed to the writing of the manuscript: CL MS EQ PK. Analysis: DH EQ. Agree with the study design, and findings: DH CL MS EQ PK. All authors have read, approved the manuscript and confirm that they meet ICMJE criteria for authorship. Trial registration: Not available. All details regarding the study can be found in the study protocol https://biblio.ugent.be/publication/8563182.

## Abstract

**Objectives:** To test authors’ experience of applying the STrengthening the Reporting of Observational Studies in Epidemiology-nutritional epidemiology (STROBE-nut) on a recently published paper. Correct understanding of the items on the reporting guidelines could lead to appropriate use.

**Design:** a cross sectional study, with a convenient sample.

**Setting:** Participants were asked to return the STROBE-nut data collection template filled on recently published papers. Next, an interview was organised to collect feedback on the process of applying the guidelines. Two researchers involved in the development of STROBE-nut completed the template using the participant’s respective published papers. The filled templates were then compared to assess the measure of agreement of the STROBE-nut items as a proxy of understanding.

**Participants:** Authors who recently published papers reporting dietary assessment or food intake data.

**Results:** We recruited 12 participants between May 2018 and June 2019. Five participants never used reporting guidelines before, while ten reported intention of future use. Half of the participants reported that the use of filling STROBE nut was useful, but some modifications are needed. Agreement between participants and experts on items reporting was generally low. Only two items had moderate weighted kappa agreement nut 1 (Kappa= 0.4, *P* 0.02) and nut 22.1 *(Kappa= 0*.*47, P* 0.01).

**Conclusions:** There is need to ensure correct understanding of STROBE-nut by authors. Revisions of STROBE-nut that make the items shorter and simpler can increase understanding. Training researchers at early stage on the aim of reporting guidelines could potentially increase proper use and decrease subjective interpretation.

## Introduction

A sizeable body of published papers does not add value to science (1). An essential effort to improve the impact of research is to ensure that all essential information is present in research manuscripts to enable correct interpretation of the study and its findings. Reporting guidelines have been developed to remind researchers to ensure essential information is included in manuscripts. Moreover, they could improve the editorial and peer review processes as they direct readers to key elements of the study. To date, over 400 reporting guidelines are available (2). Despite this however, a large proportion of researchers is still unaware of the existence and utility of reporting guidelines (3).

Efforts are needed to promote correct use of reporting guidelines in the user community i.e. authors, reviewers and editors. A few initiatives were developed to increase the use of reporting guidelines and improve their uptake including endorsement by journals. Among the initiatives are writing aids, a technological extension of a reporting guideline to increase the ease of use (4-6).

At the same time however, considerations are also needed on the side guideline developers to ensure maximum utility of reporting guidelines (7). There is a need to ensure reporting guidelines are up-to-date and provide timely recommendations (8). No previous initiative has assessed the understanding of the reporting guidelines by early career researchers. Thus, an assessment of how reporting guideline are used by authors, their perceived added value and burden is a timely effort in this regard.

STROBE-nut was developed in 2016 to improve reporting in nutrition epidemiology and dietary assessment and contains a set of 24 items (9). The guideline endorsed by various journals (10-14), provides instructions for authors to submit a checklist that indicates the page numbers where the STROBE-nut items are reporting in the manuscript. As part of the team that led the development of STROBE-nut, we monitor its use and understanding, to inform further updates and continuous improvements (15).

### Objectives and hypothesis

The objective of this study is to test authors’ experience of applying a data collection template based on the STROBE-nut table on a recently published paper and collecting their feedback on the overall experience and understanding of the items included on the checklist. We hypothesis that better understanding of the checklist’s items could lead to correct use.

## Methods and materials

### Study design, procedures and participants

We performed a cross sectional study with a convenience sample of researchers. First, we assessed understanding of the STROBE-nut items using a set of recently published papers. The authors were invited to participate by filling a data collection template based on the STROBE-nut checklist, completing the page number for each item for their paper using the information in their published papers in one of the following categories: 1-Irrelevant/not applicable 2-Fully reported on page/pages 3-Partially reported on page/pages 4-Reasons for exclusion (Annex 1), and sending it to the lead researcher of the present study. A baseline questionnaire was sent to gather participants’ characteristics (Annex 2). Second, an interview was scheduled to obtain author’s feedback on the application of the STROBE-nut on their paper in phase one (Annex 3). Third, two experts (DH and CL) who took part in the development of STROBE-nut filled the checklist using the information provided in the published paper. The purpose was to assess the agreement between the experts and the authors regarding the reporting of STROBE-nut items in the text. If the authors applied the same category where the items are in paper as the experts, this was considered as an agreement on the reporting of the item.

Participants remained in their research environment. Follow-up was conducted via online communications. The communication between the lead investigator and the participants were through email (sending the baseline questionnaire, the STROBE-nut guidelines). The interview was done over MS Skype.

Eligible criteria included authors who published papers reporting dietary assessment or food intake data. Invitations were disseminated to recruit potentially eligible authors. Ghent University academic bibliography https://biblio.ugent.be was searched using the term “nutrition” to find relevant papers. We conducted the search in May 2018. Moreover, we disseminated the invitation within our personal network and conferences including colleagues in different departments at Ghent University. Ghent University is known to have a large community of food and nutrition research, and a high Shanghai ranking in life and agriculture ranking(16).

### Pilot testing

The baseline questionnaire, the table and the Skype interview were tested with two PhD students at the UGent department of Food Technology, Safety, and Health.

### Study procedures

A baseline questionnaire was used to gather participants’ characteristics including their research experience (PhD/Post Doc), previous use of reporting guidelines, frequency of use, and motivation of use of reporting guidelines. We also assessed participant’s prior knowledge regarding reporting guidelines using a tool to assess knowledge regarding checklists (17). Subjective knowledge considering the utilization and content of the reporting guidelines was measured with two questions on a 5-point Likert scale ranging from very unknowledgeable (1) to very knowledgeable (5). Objective knowledge was measured using six true or false statements, same questions used in a previous study (6). Informed consent was collected electronically in the baseline questionnaire. Baseline information and informed consent are provided in annex 2. Participants confirmed that they have filled the baseline questionnaire during the Skype call.

### Study outcomes and measurements

Authors were asked to fill out a checklist (Annex 1, table1) and complete the page number for each item for their paper: 1-Irrelevant/not applicable 2-Fully reported on page/pages 3-Partially reported on page/pages 4-Reasons for exclusion. Example for illustration is in Annex 1.

**Table 1.**
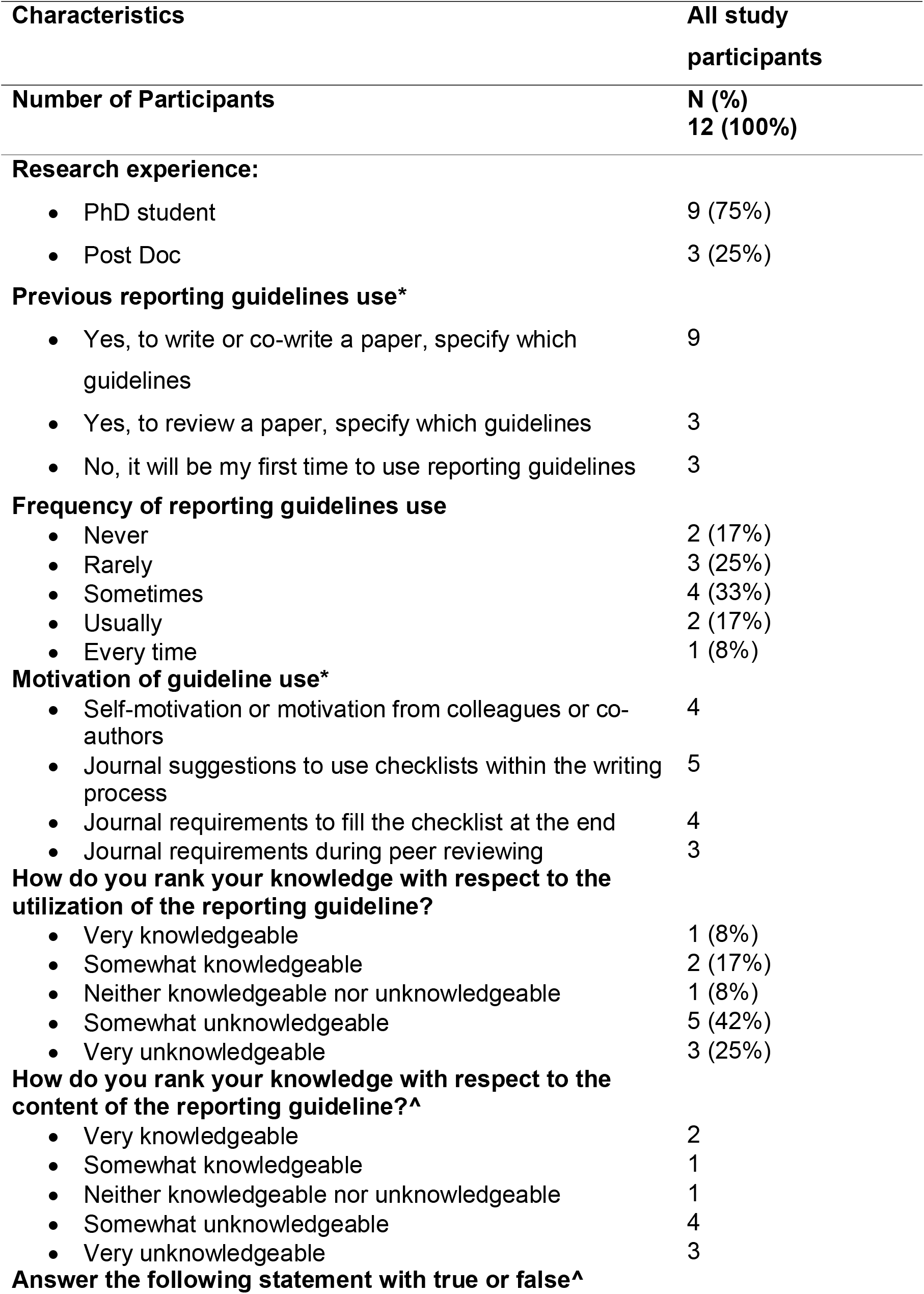
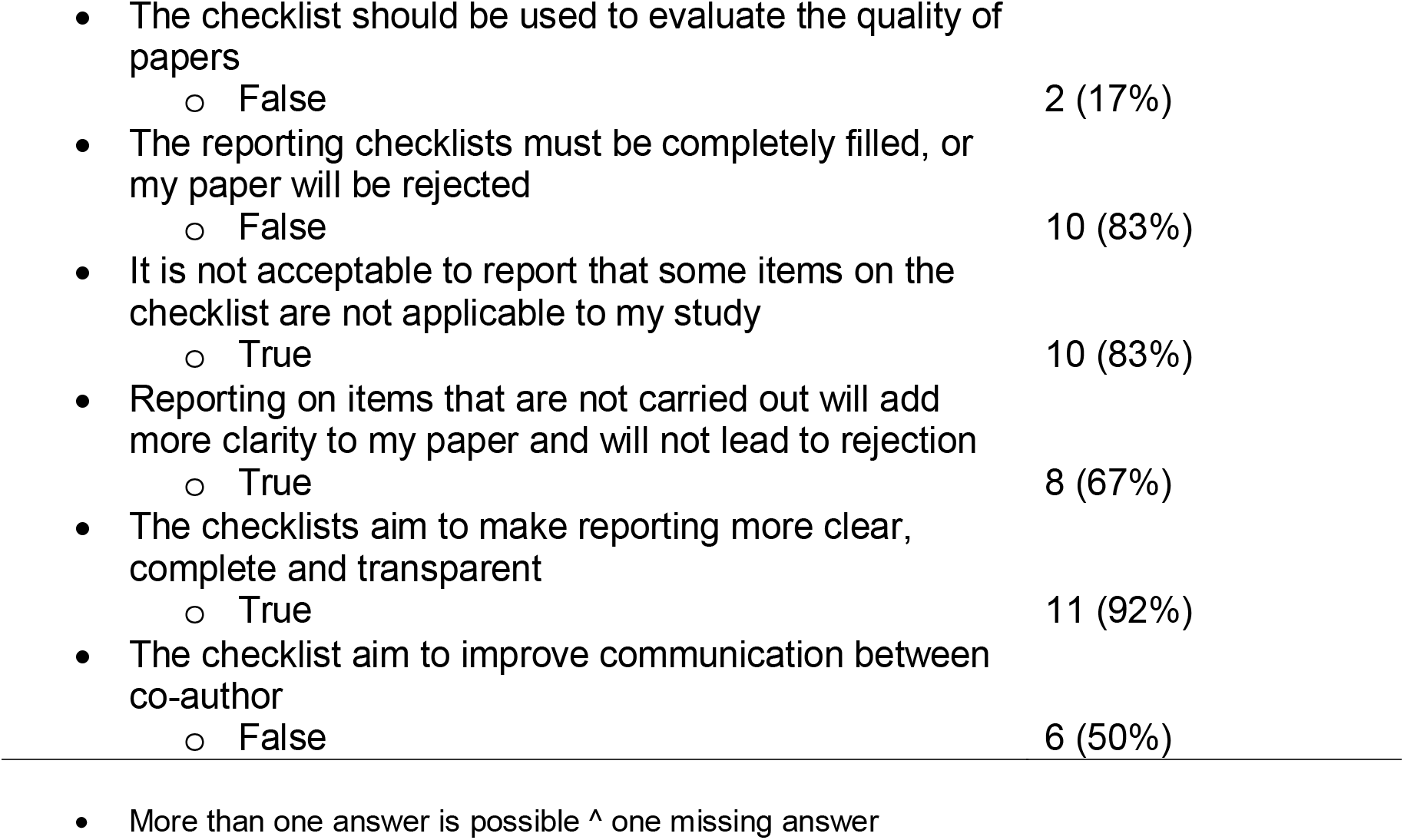
Sample characteristics.

Experts (DH and CL) only filled the first three columns: not applicable, fully reported, and partially reported on page/pages. Consensus between the experts (DH and CL) was reached through discussions for each manuscript’s items for each manuscript. The results were compared against the submitted answer for each respondent. Agreement between the author’s and the experts’ answer was calculated for every item.

After the return of the filled STROBE-nut checklist, a short semi-structured interview was conducted in English to understand the experience of the users with the reporting guidelines. The questions can be found in annex 3. We were mainly interested to have a better view on the experience of using STROBE nut, barriers, added value of and the intention for use in the next manuscript. As no previous validated questionnaire was available, we developed an interview guide for the purpose of the study. The questionnaire was pretested with two PhD students at Ghent University to assess face validity.

The interview started with a quick reminder of the study objective, intended uses of the interview results, and reassurance that the confidentiality and anonymity were protected. Permission to take notes and record the conversation electronically was requested. The Skype call took between 30 minutes and 1 hour.

### Statistical methods

Data cleaning and analysis were conducted using Stata version 14.1 (StataCorp). Descriptive statistics from the baseline questionnaire were reported using absolute numbers and percentages. For each binary question in the interview, answers were calculated, and results were reported using absolute numbers and percentages.

Agreement between participants and experts was assessed for each paper, and for each item across all papers. Three different level of agreement were calculated. 1-Agreement on filling the correct column/category (Irrelevant/not applicable, Fully reported on page/pages, Partially reported on page/pages, Reasons for exclusion.)

Cohen’s weighted kappa test was used to assess the level agreement. The following cut-offs were used: Kappa values ≤ 0 no agreement and 0.01–0.20 none to slight, 0.21–0.40 fair, 0.41– 0.60 moderate, 0.61–0.80 substantial, and 0.81–1.00 almost perfect agreement (18). The results were reported as percent agreement, and Cohen’s weighted kappa. To calculate weighted Kappa on the agreement level on the columns filled, the answers of the participants/experts were coded in ordinal order. “Not applicable” and “fully reported” were coded as 2, “partially reported” reported as 1 and “reason for exclusion” as 0. Complete reporting is regarded hence when the item is reported as fully reported or not applicable. The frequency of positive fully reported items/ not applicable were calculated for each item across studies.

### Qualitative data analysis

To extract results from interviews, anonymised answers were analysed qualitatively by two researchers (DH, EQ). DH and EQ coded all answers to each question independently. For all questions, the two researchers then discussed differences in coding per question until they reached agreement. The analysis from all questions was combined, similarities where grouped and organized in themes. Striking answers are quoted in the results to illustrate the findings. The data were anonymised after data cleaning and analysis.

### Ethics

The study was approved by the Ethics Committee of the Ghent University Hospital number EC /2018/0636. Informed consent was electronically collected and the study protocol was registered prior to the study (2^nd^ May 2018) on https://biblio.ugent.be/publication/8563182.

## Results

### Participants

We found 168 published papers after the search of Ghent University academic repository of papers on 4^th^ May 2018. The majority (n=132) papers did not report food intake data and were excluded. Finally, 3 papers were published by the same first author, 2 authors published 2 papers. Invitation emails were sent to the first authors of the remaining 32 papers to participate. Seven authors responded positively and participated in the study. Five other authors were included in the study through dissemination within personal networks. It was not possible to track response rate, as recruitment methods used personal networks, and invitations were also spread in conferences. We recruited 12 participants between May 2018 and June 2019. Data collection was completed by June 2019.

As shown in Table 1, 75% (*n = 9/12*) of the sample was PhD students. Almost half (42%) of the sample had rarely or never used any reporting guideline before considered himself or herself un-knowledgeable regarding the guidelines content or its utilization (*n = 3*). Only two (17%) participants correctly answered that reporting guidelines should not be used as an evaluation tool for the quality of the paper. Almost all participants correctly answered the three statements regarding the aim of the reporting guidelines, and the way it needs to be filled. One third stated that they sometimes used reporting guidelines.

### Skype interview results

Table 2 is a summary of the participants’ answers regarding the ease of use of a data collection template checklist. Overall, there was a positive perception of reporting guidelines by the participants. Participants expressed motivation and interest with using a reporting guidelines and/or STROBE-nut for their next manuscripts, despite certain hesitations and confusion on some sections of the data collection template.

**Table 2.**
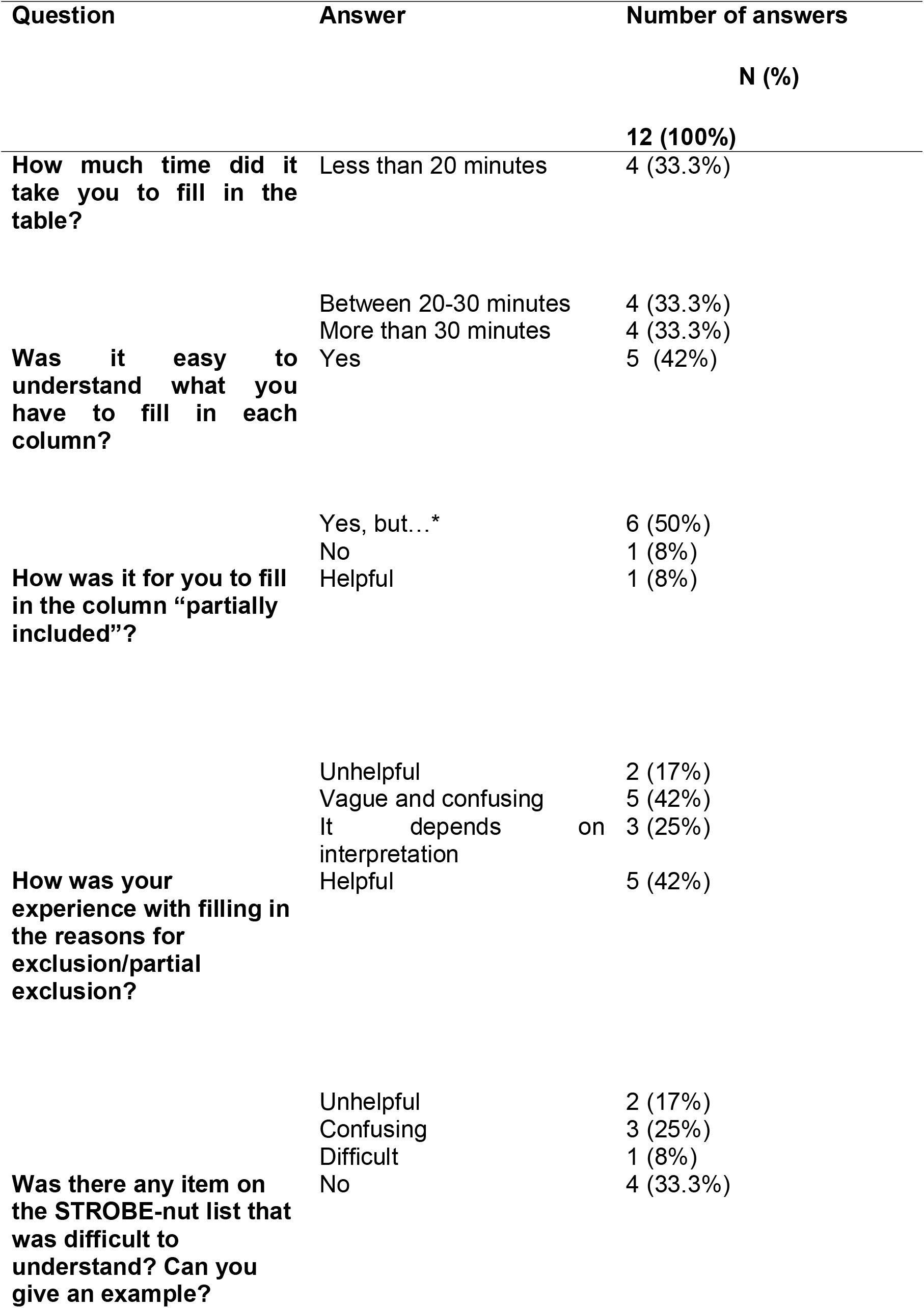
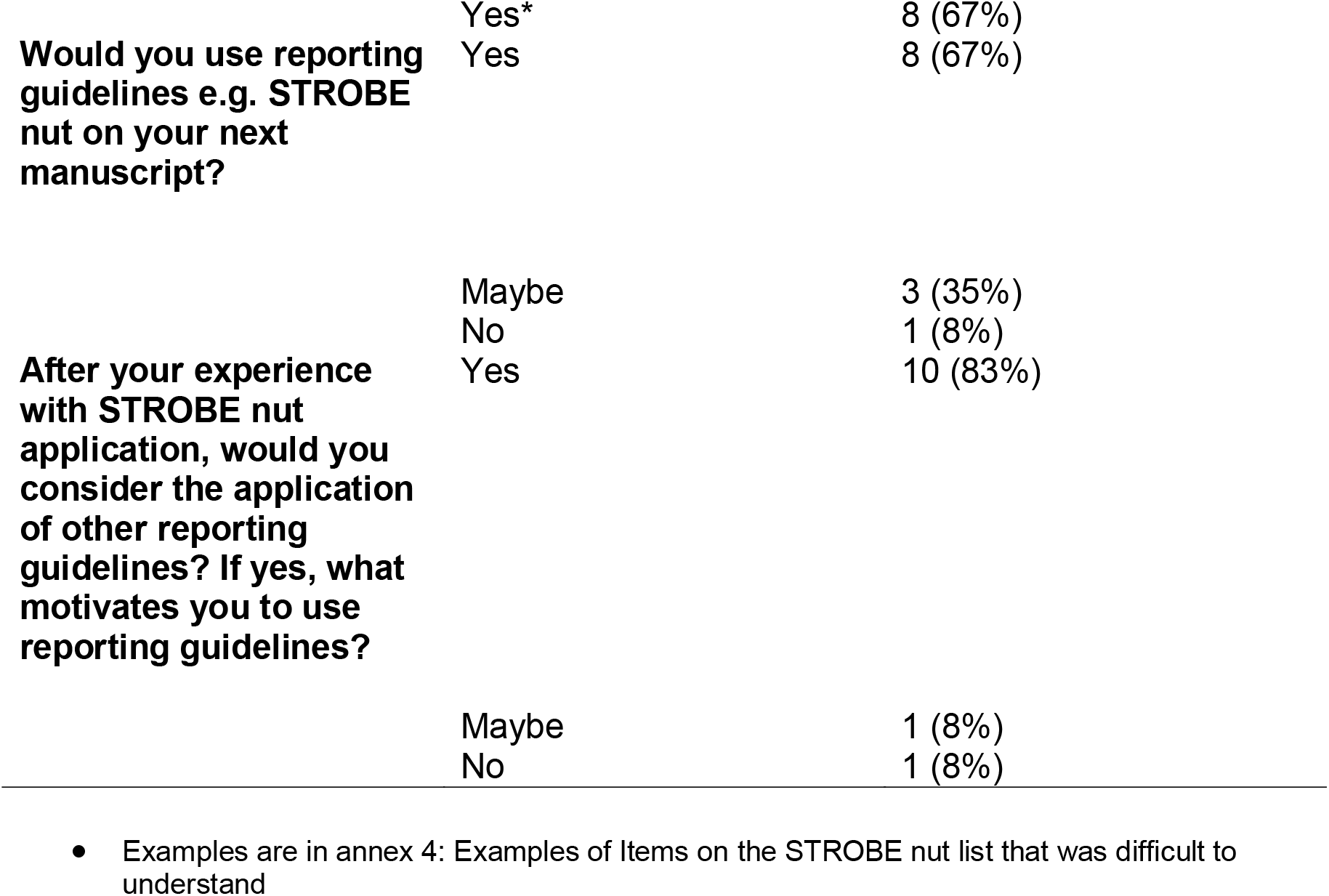
Author satisfaction regarding use of the STROBE-nut checklist Question Answer Number of answers.

Regarding the experience of filling STROBE-nut, completing the checklist was perceived fast (n= 4), easy (n=4), helpful (n=3), and comprehensive (n=2) while (n=4) authors stated that the checklist was not the most suitable for their study, as dietary assessment was not the main measurement in the study. The duration of filling the STROBE-nut checklist varied from 10 minutes to 1 hour and 55 minutes.

Applying the data collection template on already published work, made some (n=3) authors realize they missed an opportunity to include certain piece of information that could make their research more comprehensive. A participant stated having to fill the STROBE-nut checklist very quickly upon submission. When filling it again for the study, the participant realized it wasn’t filled correctly initially.

Participants perceived it challenging to apply STROBE-nut as an extension of STROBE. Authors were confused as to what extent should they fill the STROBE-nut, and if they are obliged to fill both the STROBE and the STROBE-nut extension. For example, one author said “*Yes, at first I was not hundred percent sure what the extra nutrition column meant if I had to look at the one for nutrition and the one before it*.

*But then I just interpreted it that I only use to have the nutrition one since I have a nutrition paper. I hope that it was correct otherwise it was easy. Some lines didn’t have the extra nutrition component and I still filled them, but the ones that had the nutrition component I only filled in those* related to the nutrition question.”

Participants also expressed further uncertainties on the use of STROBE-nut, i.e. when in the timeline of writing the manuscripts should STROBE-nut be applied, and reporting on all items with a limited word count. Participants also suggested introducing introduce a more user-friendly interface, such as an online system to fill in the checklist e.g. applying the checklist as an online survey during the submission process.

Generally, the authors experienced difficulties to understand the difference between “fully reported” and “partially reported” for the data collection template describing it with words like “vague, difficult and confusing’’.

Participants attributed the difficulties in filling the “fully reported” column on the checklist, to several reasons i) journal policy and word count, ii) person’s understanding of the items, the need for high level of understanding the purpose of the checklists and how to fill them iii) certain items like nut 8.1 and nut 5, nut 9 ask to report different elements in one item, however authors tend to sometimes address one element only iv) the reason why the information is excluded or partially excluded is arbitrary, subjective and depends on personal behaviour.

Authors thought that the opportunity to add reason for exclusion was quite helpful, as it helped them realize that they had missed adding available information to their manuscript: *“sometimes it was to justify why I missed it, or to understand why I missed it. It was retrospectively, so you realize maybe it was not excluded for a good reason. For me it was just to try to justify why I excluded it, it was the second analysis of the same data ‘‘*. ‘‘So *I tried to put the reason why I didn’t include it but I did not know if it was an adequate reason to exclude it or not’’*.

Participants explained that pieces of information were not reported unintentionally. They simply forgot to include it, because it was deleted by co-authors or they did not perceive it as essential content.

In addition, participants also expressed confusion on certain items, in terms of its phrasing and applicability to their respective study. These items are summarized in annex 4.

Participants were also asked to describe the barriers of using a reporting guideline. Two main themes were brought up. Firstly, making structural changes to STROBE-nut checklist itself; by breaking down the items into smaller components and providing more examples. Secondly, to offer an integrated module in academia that can make the use of reporting guideline common practice. Examples : i) teach STROBE-nut to university students, ii) create an educational module on how to correctly apply STROBE-nut, iii) integrate the guidelines into common practice such as during write up of their dissertations or written assignments.

Overall, the participants reported that applying STROBE-nut made their papers clearer and more transparent and facilitated review by others. As quoted by a participant: “*This STROBE-nut is a gold mine for me, you know if all papers report what is in STROBE-nut, it will be very easy for me to do a systematic review on nutritional topics*… *it enhances a lot of things like communication, yet this is secondary, the main part is the evidence analysis part”*.

### Inter-rater agreement results

Only two items had moderate weighted kappa agreement “nut 1 State the dietary/nutritional assessment method(s) used in the title, abstract, or keywords” (Kappa= 0.4, *P* 0.02), “nut 22 Describe the procedure for consent and study approval from ethics committee(s)” (Kappa=0.47, *P* 0.01). Ten items (nut 8.6, nut 9, nut 12.2, nut 12.3, nut 14, nut16, nut 17, nut 19, nut 20, nut 22.2 –see table 3 for full items) had none to slight agreement levels (Table 3) “nut 5 Describe any characteristics of the study settings that might affect the dietary intake or nutritional status of the participants, if applicable” and “nut-8.2 Describe and justify food composition data used. Explain the procedure to match food composition” with had zero agreement. Nine items (nut 6, nut 7.1, nut 7.2, nut 8.1, nut 8.3, nut 8.5, nut 11, nut 12.1, nut 13) yielded negative Kappa indicating agreement worse than expected and systematic disagreement (18). Agreement on the overall reporting per study between the author and experts is reported in table 4. Average agreement across all papers was 48.9%. The Frequency of positive fully reported items/ not applicable varied between 100% (nut 8.4) and 17% (nut 6, nut 8.6 and nut 22.2). Nut item 9 and 17 were both zero (Table 5).

**Table 3.** Agreement per item between experts and researchers.

| STROBE nut items | %<br>Classification<br>agreement | Weighted<br>Kappa | P-value |
| --- | --- | --- | --- |
| **Strobe nut-1** “State the dietary/nutritional assessment method(s) used in the title, abstract, or keywords.” | 79.2% | 0.44 | 0.02 |
| **Strobe nut 5** “Describe any characteristics of the study settings that might affect the dietary intake or nutritional status of the participants, if applicable” | 58.3% | 0.00 | 0.5 |
| **Strobe nut-6** “Report particular dietary, physiological or nutritional characteristics that were considered when selecting the target population.” | 37.5% | -0.30 | 0.95 |
| **Strobe nut-7.1** “Clearly define foods, food groups, nutrients, or other food components.” | 70.8% | -0.24 | 0.86 |
| **Strobe nut-7.2** “When using dietary patterns or indices, describe the methods to obtain them and their nutritional properties.” | 75% | -0.13 | 0.68 |
| **Strobe nut-8.1** “Describe the dietary assessment method(s), e.g., portion size estimation, number of days and items recorded, how it was developed and administered, and how quality was assured. Report if and how supplement intake was assessed.” | 58.3% | -0.15 | 0.80 |
| **Strobe nut-8.2** “Describe and justify food composition data used. Explain the procedure to match food composition with consumption data. Describe the use of conversion factors, if applicable.” | 70.8% | 0.00 | 0.5 |
| **Strobe nut-8.3** “Describe the nutrient requirements, recommendations, or dietary guidelines and the evaluation approach used to compare intake with the dietary reference values, if applicable.” | 79.2% | -0.15 | 0.76 |
| **Strobe nut 8.4\*** “When using nutritional biomarkers, additionally use the STROBE Extension for Molecular Epidemiology | - | - | - |
| (STROBE-ME). Report the type of biomarkers used and their usefulness as dietary exposure markers” |  |  |  |
| **Strobe nut-8.5** “Describe the assessment of nondietary data (e.g., nutritional status and influencing factors) and timing of the assessment of these variables in relation to dietary assessment.” | 58.3% | -0.07 | 0.68 |
| **Strobe nut-8.6** “Report on the validity of the dietary or nutritional assessment methods and any internal or external validation used in the study, if applicable.” | 36.4% | 0.01 | 0.46 |
| **Strobe nut-9** “Report how bias in dietary or nutritional assessment was addressed, e.g., misreporting, changes in habits as a result of being measured, or data imputation from other sources” | 50% | 0.1 | 0.17 |
| **Strobe nut 11** “Explain categorization of dietary/nutritional data (e.g., use of N-tiles and handling of nonconsumers) and the choice of reference category, if applicable.” | 72.7% | -0.14 | 0.69 |
| **Strobe nut 12.1** “Describe any statistical method used to combine dietary or nutritional data, if applicable.” | 79.2% | -0.11 | 0.73 |
| **Strobe nut 12.2** “Describe and justify the method for energy adjustments, intake modeling, and use of weighting factors, if applicable.” | 58.3% | 0.03 | 0.42 |
| **Strobe nut 12.3** “Report any adjustments for measurement error, i.e. from a validity or calibration study.” | 36.4% | 0.10 | 0.23 |
| **Strobe nut-13** “Report the number of individuals excluded based on missing, incomplete or implausible dietary/nutritional data” | 36.4% | -0.09 | 0.67 |
| **Strobe nut-14** “Give the distribution of participant characteristics across the exposure variables if applicable. Specify if food | 72.7% | 0.08 | 0.33 |
| consumption of total population or consumers only were used to obtain results.” |  |  |  |
| **Strobe nut-16** “Specify if nutrient intakes are reported with or without inclusion of dietary supplement intake, if applicable.” | 45.0% | 0.08 | 0.33 |
| **Strobe nut-17** “Report any sensitivity analysis (e.g., exclusion of misreporters or outliers) and data imputation, if applicable.” | 33.3% | 0.03 | 0.27 |
| **Strobe nut-19** “Describe the main limitations of the data sources and assessment methods used and implications for the interpretation of the findings.” | 70.8% | 0.11 | 0.31 |
| **Strobe nut-20** “Report the nutritional relevance of the findings, given the complexity of diet or nutrition as an exposure.” | 66.7% | 0.14 | 0.20 |
| **Strobe nut-22.1** “Describe the procedure for consent and study approval from ethics committee(s).” | 87.5% | 0.47 | 0.01 |
| **Strobe nut-22.2** “Provide data collection tools and data as online material or explain how they can be accessed.” | 54.5% | 0.13 | 0.28 |

\*Item nut 8.4 did not have any response, as it was not applicable for any of the included studies
\*(Important to note that the opinions are based from the use of a data collection template)

**Table 4.** Average agreements on reporting of STROBE-nu items per paper between experts and researchers.

| **Paper** | **%**<br>**classification**<br>**agreement** | **Kappa** | **P-value** |
| --- | --- | --- | --- |
| Paper 1 | 42.9% | -0.10 | 0.72 |
| Paper 2 | 61.1% | 0 | 0.5 |
| Paper 3 | 50.0% | 0.16 | 0.07 |
| Paper 4 | 54.1% | 0.22 | 0.05 |
| Paper 5 | 58.3% | 0.29 | 0.01 |
| Paper 6 | 37.5% | 0.05 | 0.34 |
| Paper 7 | 45.8% | -0.06 | 0.67 |
| Paper 8 | 45.8% | 0.16 | 0.12 |
| Paper 9 | 45.8% | 0.14 | 0.13 |
| Paper 10 | 41.7% | 0.07 | 0.10 |
| Paper 11 | 66.7% | 0.19 | 0.09 |
| Paper 12 | 37.5% | -0.18 | 0.88 |
| Average agreement across papers | 48.9% |  |  |

**Table 5.** Frequency of positive fully reported items/ not applicable.

| **STROBE nut items** | **Frequency of positive fully reported items/ not applicable 12 (100%)** |
| --- | --- |
| **Strobe nut-1** “State the dietary/nutritional assessment method(s) used in the title, abstract, or keywords.” | 8 (67%) |
| **Strobe nut 5** “Describe any characteristics of the study settings that might affect the dietary intake or nutritional status of the participants, if applicable” | 3 (25%) |
| **Strobe nut-6** “Report particular dietary, physiological or nutritional characteristics that were considered when selecting the target population.” | 2 (17%) |
| **Strobe nut-7.1** “Clearly define foods, food groups, nutrients, or other food components.” | 7 (58%) |
| **Strobe nut-7.2** “When using dietary patterns or indices, describe the methods to obtain them and their nutritional properties.” | 11 (92%) |
| **Strobe nut-8.1** “Describe the dietary assessment method(s), e.g., portion size estimation, number of days and items recorded, how it was developed and administered, and how quality was assured. Report if and how supplement intake was assessed.” | 4 (33%) |
| **Strobe nut-8.2** “Describe and justify food composition data used. Explain the procedure to match food composition with consumption data. Describe the use of conversion factors, if applicable.” | 6 (50%) |
| **Strobe nut-8.3** “Describe the nutrient requirements, recommendations, or dietary guidelines and the evaluation approach used to compare intake with the dietary reference values, if applicable.” | 9 (75%) |
| **Strobe nut 8.4\*** “When using nutritional biomarkers, additionally use the STROBE Extension for Molecular Epidemiology (STROBE-ME). Report the type of biomarkers used and their usefulness as dietary exposure markers” | 12 (100%) |
| **Strobe nut-8.5** “Describe the assessment of nondietary data (e.g., nutritional status and influencing factors) and timing of the assessment of these variables in relation to dietary assessment.” | 3 (25%) |
| **Strobe nut-8.6** “Report on the validity of the dietary or nutritional assessment methods and any internal or external validation used in the study, if applicable.” | 2 (17%) |
| **Strobe nut-9** “Report how bias in dietary or nutritional assessment was addressed, e.g., misreporting, changes in habits as a result of being measured, or data imputation from other sources” | 0 (0%) |
| **Strobe nut 11** “Explain categorization of dietary/nutritional data (e.g., use of N-tiles and handling of nonconsumers) and the choice of reference category, if applicable.” | 11 (92%) |
| **Strobe nut 12.1** “Describe any statistical method used to | 9 (75%) |
| combine dietary or nutritional data, if applicable.” |  |
| **Strobe nut 12.2** “Describe and justify the method for energy adjustments, intake modeling, and use of weighting factors, if applicable.” | 6 (50%) |
| **Strobe nut 12.3** “Report any adjustments for measurement error, i.e. from a validity or calibration study.” | 3 (25%) |
| **Strobe nut-13** “Report the number of individuals excluded based on missing, incomplete or implausible dietary/nutritional data” | 4 (33%) |
| **Strobe nut-14** “Give the distribution of participant characteristics across the exposure variables if applicable. Specify if food consumption of total population or consumers only were used to obtain results.” | 7 (58%) |
| **Strobe nut-16** “Specify if nutrient intakes are reported with or without inclusion of dietary supplement intake, if applicable.” | 3 (25%) |
| **Strobe nut-17** “Report any sensitivity analysis (e.g., exclusion of misreporters or outliers) and data imputation, if applicable.” | 0 (0%) |
| **Strobe nut-19** “Describe the main limitations of the data sources and assessment methods used and implications for the interpretation of the findings.” | 6 (50%) |
| **Strobe nut-20** “Report the nutritional relevance of the findings, | 5 (42%) |

| STROBE nut items | Frequency of<br>positive fully<br>reported items/ not<br>applicable 12 (100%) |
| --- | --- |
| given the complexity of diet or nutrition as an exposure.” |  |
| **Strobe nut-22.1** “Describe the procedure for consent and study approval from ethics committee(s).” | 8 (67%) |
| **Strobe nut-22.2** “Provide data collection tools and data as online material or explain how they can be accessed.” | 2 (17%) |

## Discussion

This study assessed user experience with the use of STROBE-nut reporting guidelines in nutrition research and agreement with expert classification. Participants in this study seem to understand the added value of using STROBE-nut when reporting their studies and are willing to use it. Nevertheless, they faced many obstacles applying STROBE-nut on their published papers, including the difficulty to apply STROBE-nut as an extension of STROBE. Updates that make the items shorter and simpler can reduce subjectivity according to authors’ feedback, as well as integrating reporting guidelines in the learning process at earlier stage e.g. Bachelors or Masters degrees could increase the levels of understanding.

To our knowledge, our study was the first to assess the user’s understanding from the actual application of reporting guidelines to recently published papers. Suggestions and feedback from our qualitative review provided valuable insight on how authors perceive the items, and how to update the STROBE-nut to suit their needs.

Looking at the process of updating 7 other reporting guidelines within the last ten years (19-22) (23-25). Only The Consolidated Standards Of Reporting Trials “CONSORT” Statement for Randomized Trials of Nonpharmacologic Treatments “CONSORT-NPT 2017” (20) has identified authors who used CONSORT-NPT - asking for suggestions and feedback per each item on the checklist.

Most of our participants are PhD students, and almost half of the sample rarely or never used reporting guidelines before. The application of a STROBE-nut table by researchers on their published manuscript has potentially increase their intention to use the tool in further writing exercises. The observed high intention of future use is similar to result we have obtained studying the use reporting guidelines as writing aid (6).

As the application and endorsement of STROBE-nut is increasing, there is a need to ensure it is used correctly. Participants suggested developing an education module on STROBE-nut. Thus we have created a module which is shared on STROBE-nut website http://www.strobe-nut.org/content/strobe-nut-lecture. We are currently piloting it and welcome contributions. Also, participants suggested applying the checklist as an online survey within the submission process. This is in line with a few initiatives that are working on improving adherence to reporting guidelines including integrating them in the writing process(6, 26).

Our study has several strengths including the application of STROBE-nut on recently published papers and collecting users’ feedback that could be integrated in further updates. It provides insights into the behaviour of filling out a checklist. The study also served as a learning experience for the participants. Doing an exercise like this is beneficial and can be seen as an interactive learning experience. On the other hand, the study only involved a small sample size, smaller than what was indicated in the protocol (27). This could be due to limited search using Ghent University academic bibliography and studies reporting food intake data and dietary assessment within the timeframe of the study. Kappa values are affected by the prevalence of the incidence under consideration, thus the small sample size could possible resulted in lower Kappa levels (28).

Finally, this study is an reminder to be vigilant about the added value and potential burden due to the introduction of reporting guidelines on researchers (29). To ensure optimal application and timely revisions of reporting guideline, it is essential to consider the primary target audience: authors of research manuscripts during the development and modifications of reporting guidelines.

## Supporting information

Annex 1

Annex 2

Annex 3

Annex 4

Annex 5

## Data Availability

Anonymised data is added as Annex 5.

## Acknowledgments

The authors Dr. Nathalie De Cock and Dr. Abdulhalik Workicho, for their input during pilot testing of the questionnaires. Alemayehu Argaw for his input on the interpretation of Kappa test.

## Abbreviations

EQUATOR: The Enhancing the QUAlity and Transparency of health Research
CONSORT: The Consolidated Standards Of Reporting Trials
CONSORT-NPT 2017: The Consolidated Standards Of Reporting Trials Statement for Randomized Trials of Nonpharmacologic Treatments
PRISMA: the Preferred Reporting Items for Systematic Reviews and Meta-Analyses Statement
STROBE: the Strengthening the Reporting of Observational Studies in Epidemiology
Statement, and “STROBE-nut: the STrengthening the Reporting of Observational Studies in Epidemiology-nutritional epidemiology statement

