## Supplementary material for "Assessment of authors understanding of the STROBE-nut reporting guidelines": Annex 1

Lachat C et al. (2016) STrengthening the Reporting of OBservational studies in Epidemiology – Nutritional Epidemiology (STROBE-nut): an extension of the STROBE statement. Plos Medicine 13(6) <http://dx.doi.org/10.1371/journal.pmed.1002036> [pdf](http://journals.plos.org/plosmedicine/article/asset?id=10.1371%2Fjournal.pmed.1002036.PDF) or [online](http://journals.plos.org/plosmedicine/article?id=10.1371/journal.pmed.1002036) version.

| **Item** | **Item nr** | **STROBE recommendations** | **Extension for Nutritional Epidemiology studies (STROBE-nut)** | **NA** | **Fully Reported on page #** | **Partially Reported on page #** | **Reasons for exclusion** |
| --- | --- | --- | --- | --- | --- | --- | --- |
| **Title and**  **Abstract** | 1 | (a) Indicate the study’s design with a commonly used term in the title or the abstract. | **nut-1** State the dietary/nutritional assessment method(s) used in the title, abstract, or keywords. |  |  |  |  |
|  |  | (b) Provide in the abstract an informative and balanced summary of what was done and what was found. |  |  |  |  |  |
| **Introduction** |  |  |  |  |  |  |  |
| Background rationale | 2 | Explain the scientific background and rationale for the investigation being reported. |  |  |  |  |  |
| Objectives | 3 | State specific objectives, including any pre-specified hypotheses. |  |  |  |  |  |
| **Methods** |  |  |  |  |  |  |  |
| Study design | 4 | Present key elements of study design early in the paper. |  |  |  |  |  |
| Settings | 5 | Describe the setting, locations, and relevant dates, including periods of recruitment, exposure, follow-up, and data collection. | **nut-5** Describe any characteristics of the study settings that might affect the dietary intake or nutritional status of the participants, if applicable. |  |  |  |  |
| Participants | 6 | a) Cohort study—Give the eligibility criteria, and the sources and methods of selection of participants. Describe methods of follow-up.  Case-control study—Give the eligibility criteria, and the sources and methods of case ascertainment and control selection. Give the rationale for the choice of cases and controls.  Cross-sectional study—Give the eligibility criteria, and the sources and methods of selection of participants.  (b) Cohort study—For matched studies, give matching criteria and number of exposed and unexposed.  Case-control study—For matched studies, give matching criteria and the number of controls per case. | **nut-6** Report particular dietary, physiological or nutritional characteristics that were considered when selecting the target population. |  |  |  |  |
| Variables | 7 | Clearly define all outcomes, exposures, predictors, potential confounders, and effect modifiers. Give diagnostic criteria, if applicable. | **nut-7.1** Clearly define foods, food groups, nutrients, or other food components. |  |  |  |  |
|  |  |  | **nut-7.2** When using dietary patterns or indices, describe the methods to obtain them and their nutritional properties. |  |  |  |  |
| Data sources - measurements | 8 | For each variable of interest, give sources of data and details of methods of assessment (measurement).Describe comparability of assessment methods if there is more than one group. | **nut-8.1** Describe the dietary assessment method(s), e.g., portion size estimation, number of days and items recorded, how it was developed and administered, and how quality was assured. Report if and how supplement intake was assessed. |  |  |  |  |
|  |  |  | **nut-8.2** Describe and justify food composition data used. Explain the procedure to match food composition with consumption data. Describe the use of conversion factors, if applicable. |  |  |  |  |
|  |  |  | **nut-8.3** Describe the nutrient requirements, recommendations, or dietary guidelines and the evaluation approach used to compare intake with the dietary reference values, if applicable. |  |  |  |  |
|  |  |  | **nut-8.4** When using nutritional biomarkers, additionally use the STROBE Extension for Molecular Epidemiology (STROBE-ME). Report the type of biomarkers used and their usefulness as dietary exposure markers. |  |  |  |  |
|  |  |  | **nut-8.5** Describe the assessment of nondietary data (e.g., nutritional status and influencing factors) and timing of the assessment of these variables in relation to dietary assessment. |  |  |  |  |
|  |  |  | **nut-8.6** Report on the validity of the dietary or nutritional assessment methods and any internal or external validation used in the study, if applicable. |  |  |  |  |
| Bias | 9 | Describe any efforts to address potential sources of bias. | **nut-9** Report how bias in dietary or nutritional assessment was addressed, e.g., misreporting, changes in habits as a result of being measured, or data imputation from other sources |  |  |  |  |
| Study Size | 10 | Explain how the study size was arrived at. |  |  |  |  |  |
| Quantitative variables | 11 | Explain how quantitative variables were handled in the analyses. If applicable, describe which groupings were chosen and why. | **nut-11** Explain categorization of dietary/nutritional data (e.g., use of N-tiles and handling of nonconsumers) and the choice of reference category, if applicable. |  |  |  |  |
| Statistical  Methods | 12 | (a) Describe all statistical methods, including those used to control for confounding  (b) Describe any methods used to examine subgroups and interactions.  (c) Explain how missing data were addressed.  (d) Cohort study—If applicable, explain how loss to follow-up was addressed.  Case-control study—If applicable, explain how matching of cases and controls was addressed.  Cross-sectional study—If applicable, describe analytical methods taking account of sampling strategy.  (e) Describe any sensitivity analyses. | **nut-12.1** Describe any statistical method used to combine dietary or nutritional data, if applicable. |  |  |  |  |
|  |  |  | **nut-12.2** Describe and justify the method for energy adjustments, intake modeling, and use of weighting factors, if applicable. |  |  |  |  |
|  |  |  | **nut-12.3** Report any adjustments for measurement error, i.e,. from a validity or calibration study. |  |  |  |  |
| **Results** |  |  |  |  |  |  |  |
| Participants | 13 | (a) Report the numbers of individuals at each stage of the study—e.g., numbers potentially eligible, examined for eligibility, confirmed eligible, included in the study, completing follow-up, and analyzed.  (b) Give reasons for non-participation at each stage.  (c) Consider use of a flow diagram. | **nut-13** Report the number of individuals excluded based on missing, incomplete or implausible dietary/nutritional data. |  |  |  |  |
| Descriptive data | 14 | (a) Give characteristics of study participants (e.g., demographic, clinical, social) and information on exposures and potential confounders  (b) Indicate the number of participants with missing data for each variable of interest  (c) Cohort study—Summarize follow-up time (e.g., average and total amount) | **nut-14** Give the distribution of participant characteristics across the exposure variables if applicable. Specify if food consumption of total population or consumers only were used to obtain results. |  |  |  |  |
| Outcome data | 15 | Cohort study—Report numbers of outcome events or summary measures over time.  Case-control study—Report numbers in each exposure category, or summary measures of exposure.  Cross-sectional study—Report numbers of outcome events or summary measures. |  |  |  |  |  |
| Main results | 16 | (a) Give unadjusted estimates and, if applicable, confounder-adjusted estimates and their precision (e.g., 95% confidence interval).  Make clear which confounders were adjusted for and why they were included.  (b) Report category boundaries when continuous variables were categorized.  (c) If relevant, consider translating estimates of relative risk into absolute risk for a meaningful time period. | **nut-16** Specify if nutrient intakes are reported with or without inclusion of dietary supplement intake, if applicable. |  |  |  |  |
| Other analyses | 17 | Report other analyses done—e.g., analyses of subgroups and interactions and sensitivity analyses. | **nut-17** Report any sensitivity analysis (e.g., exclusion of misreporters or outliers) and data imputation, if applicable. |  |  |  |  |
| **Discussion** |  |  |  |  |  |  |  |
| Key results | 18 | Summarize key results with reference to study objectives. |  |  |  |  |  |
| Limitation | 19 | Discuss limitations of the study, taking into account sources of potential bias or imprecision. Discuss both direction and magnitude of any potential bias. | **nut-19** Describe the main limitations of the data sources and assessment methods used and implications for the interpretation of the findings. |  |  |  |  |
| Interpretation | 20 | Give a cautious overall interpretation of results considering objectives, limitations, multiplicity of analyses, results from similar studies, and other relevant evidence. | **nut-20** Report the nutritional relevance of the findings, given the complexity of diet or nutrition as an exposure. |  |  |  |  |
| Generalizability | 21 | Discuss the generalizability (external validity) of the study results. |  |  |  |  |  |
| **Other information** |  |  |  |  |  |  |  |
| Funding | 22 | Give the source of funding and the role of the funders for the present study and, if applicable, for the original study on which the present article is based. |  |  |  |  |  |
| *Ethics* |  |  | **nut-22.1** Describe the procedure for consent and study approval from ethics committee(s). |  |  |  |  |
| *Supplementary material* |  |  | **nut-22.2** Provide data collection tools and data as online material or explain how they can be accessed. |  |  |  |  |
