## Supplementary material for "Assessment of authors understanding of the STROBE-nut reporting guidelines": Annex 2

Application of Strengthening The Reporting of OBservational Studies in Epidemiology – Nutritional Epidemiology “STROBE nut” on recently published manuscripts to assess user experience and increase adherence to reporting guidelines: Cross Sectional Study

Coordinating Investigator:	Prof. Carl Lachat

Principal Investigator:	Dana Hawwash

Dear Researcher,

You are invited to participate in a study to test the application of STROBE nut during the writing of a scientific manuscript related to nutrition. Before you decide to participate in this study, it is good to read this form as it explains the study clearly and states your rights and our responsibilities.

**Purpose and description of the study**

We have recently developed STROBE nut^[1]^, which is a checklist of 24 relevant items in addition to Strengthening The Reporting of OBservational Studies in Epidemiology “STROBE” checklist that need to be reported in a nutrition manuscript when submitted for publication. The reporting guidelines aim to increase transparency and completeness of reporting. It is expected that the application of STROBE nut will support the completeness of the reporting of nutrition paper, yet we need to understand authors’ experiences with the checklist, and make the necessary modifications to satisfy the needs of the authors, and increase the checklist applicability.

**Objective of the study**

The objective is to test authors experience regarding the application of STROBE nut table. We will ask the participants to familiarize themselves with the STROBE nut items, by applying a modified version on their recently published nutrition manuscript, and collect the filled table and their feedback through a Skype interview.

The reason of applying the reporting guidelines on a recently published paper is to give the authors an opportunity to be familiar with the checklist items in a relaxed setting (no pressure of publishing), on a familiar topic. The idea is to practice self-judgment on whether using such guidance could improve their manuscripts scientific content or not.

**How the study is done**

To evaluate the objectives a cross-sectional study with convenience sampling is set up. The participants will stay in their research/ work environment.

Upon acceptance to participate, baseline information will be collected using online questionnaire; you will be given the STROBE nut modified checklist layout under the form of a Microsoft Word document (the table has four columns to fill: NA, fully reported on page, partially reported on page #, reasons for exclusion or partial exclusion). You will be asked to go through your recently published manuscript and fill the modified table for all items.

**Example**

STROBE **nut-8.1** “Describe the dietary assessment method(s), e.g., portion size estimation, number of days and items recorded, how it was developed and administered, and how quality was assured. Report if and how supplement intake was assessed.”

How to fill the table?

**NA column**

1. In this study, you need to state if this item is not applicable in your published manuscript.

**Fully reported on page #** **column**

2. If it is applicable and it is fully reported, then you are asked to state the number of the page/s where the information is in the manuscript in the second column of the MS word table.

**Partially reported on page #** **column**

3. We have added a special column for the purpose of this study called “Partially reported on page #” Since an item can ask for more than one piece of information, of which you could have reported on one or two but not all. If for instance, in the given example STROBE nut-8.1 you only report on portion size estimation, number of days and items recorded without reporting on how the tool was developed and administered, or how the quality was assured, then you need to explain reasons for partial reporting in the third column. Partial reporting can have many reasons including, the information is not relevant for the study, overlooking the issue, having it done in another study, or having done it yet not explicitly stating it in the text.

**Reasons for exclusion or partial exclusion** **column**

4. If the item is applicable yet you haven’t reported on it in the published manuscript, or you have it partially reported then you could state reasons for exclusion. When you provide information on the reasons for exclusion, it increases transparency and clarity. It is also part of good research practices, where authors can learn how pervious research was done.

You need to go through the STROBE and STROBE nut items one by one and fill it in the same way explained above. You will be given a period of a week (It is tested: filling the table takes between 15-60 minutes of your time). After this exercise, you will be asked to schedule a Skype interview (30-45 minutes) to provide us with feedback based on your experience filling in the table.

The study consists of 3 steps:

1- Filling a 3 minutes baseline questionnaire, and providing the informed consent (online via email communication)
2- Filling and sending back the MS Table with number of page/s where the information are and providing comments when the item is not included within a period of one week. Two researchers DH and CL as external experts will also fill the table for each participant’s publication simultaneously.
3- Half an hour Skype call scheduled with study organizer based on your availability to provide insight on your experience filling in the table

**Voluntary participation**

Your participation in this study is entirely voluntarily. You have the right to refuse to participate in the study without explanation. You also have the right to stop your participation in the study at any time, even if you have signed this informed consent form.

**inconveniences**

The study will require some time investment from your end (one hour maximum filling the table and half an hour Skype call), and the application of the reporting guidelines on a paper that has already been approved and published.

The Skype call will be recorded. The data will be saved and stored until the paper is published and then the data will be discarded

**Benefits**

You will familiarize yourself with STROBE nut reporting guideline and choose for yourself to apply it on your next manuscript.

If you decide on using STROBE nut more often and you need feedback, you can schedule a Skype call with Dana Hawwash to provide further guidance.

Your effort will be acknowledged in the manuscript upon your consensus.

**Protection of your private life**

Your identity and your participation in this study will be treated strictly confidential. The specific information we obtain from you (email address, Skype recording and the filled in table with the reasons of exclusion) will not be shared with anybody, except the study investigators. Reasons for exclusion are used in this study to give insight on the STROBE nut applicability and not on your ability to carry high quality research. All data will be coded by using participant unique identity numbers. Your name will not appear in any reports or publication resulting from this study. After the study is completed, you may request information about the study results.

**Ethics committee**

The study was presented to the Ethics Committee of the University Hospital in Ghent for review.. The study was approved by the commission number EC/2018/0636

**Contact persons in case you have questions about this study**

If you have any questions concerning your participation in this study, you can always contact or the Project coordinator Prof. dr. Carl Lachat of the PhD, at +03292649377 of the Department of Food Technology, Safety and Health, Faculty of Bioscience Engineering of Ghent University, Belgium

### Baseline questionnaire

**Dear researcher**

Thank you for accepting our invitation to participate in our study. Before the start of the trial, please complete this baseline questionnaire. The questionnaire should not take more than 5 minutes of your time.

Informed Consent

- I declare that I have been informed about the purpose of this study and understand that I can refuse to answer a particular question and withdraw when I like. My name won’t be associated in any publication with the collected information. I accept that there is neither remuneration nor direct benefit for me.

### General information

**Before filling the questionnaire, please provide the following details**

Full name:

Email:

Picked number:

The current working title of the paper ( we understand that title can be modified at a later stage)

Research experience:

-PhD student

-Post Doc

-Professor

**Q1 Have you used a reporting guideline like PRISMA, CONSORT or STROBE before? (Tick all those that apply)**

- Yes, to write or co-write a paper (1), specify which guidelines
- Yes, to review a paper (2), specify which guidelines
- No, it will be my first time to use reporting guidelines (3)

**If answer is yes to the above question, then this question will show up**

In General, how often do you use reporting guidelines?

Never	Rarely	Sometimes	Usually	Every time

**Q2 What motivated you to use the guideline?**

- Self motivation or motivation from colleagues or co-authors
- Journal suggestions to use checklists within the writing process
- Journal requirements to fill the checklist at the end
- Journal requirements during peer reviewing

**Subjective knowledge**

**The following questions only apply to PRISMA, CONSORT, STROBE and STROBE nut**

**Q3 A) How do you rank your knowledge with respect to the utilization of the reporting guideline?**

- Very knowledgeable
- Somewhat knowledgeable
- Neither knowledgeable nor unknowledgeable
- Somewhat knowledgeable
- Very unknowledgeable

**Q3 B) how do you rank your knowledge with respect to the content of the reporting guideline?**

- Very knowledgeable
- Somewhat knowledgeable
- Neither knowledgeable nor unknowledgeable
- Somewhat knowledgeable
- Very unknowledgeable

**Objective knowledge**

**The following questions only apply to PRISMA, CONSORT, STROBE and STROBE nut**

**Q4 Answer the following statement with true or false**

- The reporting guidelines should be used to evaluate the quality of papers
- The reporting guideline must be completely filled with existing information in my paper, or my paper will be rejected
- It is not acceptable to report that some items on the checklist are not applicable to my study
- Reporting on items that are not carried out will add more clarity to my paper and will not lead to rejection
- The reporting guidelines aim to make reporting more clear, complete and transparent
- Reporting guidelines were developed to improve communication between the co-authors

**Informed consent form**

Before you agree to participate in this study, you need to be aware that:

- The study was presented to the Ethics Committee of the University Hospital in Ghent for review. The study was approved by the commission number EC/2018/0636
- This clearance is not to be taken as an obligation to take part in this study.
- This study is conducted according to guidelines for good clinical practice (ICH / GCP) and the Helsinki Declaration (version 2013) designed to protect people participating in clinical studies. In no case you should consider the approval by the ethics commission as an encouragement to participate in this study.
- Your participation is only voluntary. If you wish, you can withdraw from this study at any point, even after providing consent. You can withdraw by contacting the researchers through email or telephone. You do not have to motivate or explain the decision of withdrawal. Your data will be discarded and not be used in the analysis
- You can always revise your answers when filling the forms
- Your data will be coded and saved as such. Researchers not involved in the data collection will not have access to your personal data and name.
- You can contact the researcher or the coordinator of the project at any time if you wish to obtain more information regarding this study.

I, undersigned, declare that I have been informed about the purpose of this study and understand that I can refuse to answer a particular question and withdraw when I like. My name won’t be associated in any publication with the collected information. I accept that there is neither remuneration nor direct benefit for me.

My consent will be confirmed by clicking this link to the baseline online questionnaire

| Principal Investigator  Dana Hawwash  MSc, Department of Food Technology,  Safety and Health, Faculty of Bioscience Engineering | Project coordinator  Prof. dr. Carl Lachat  PhD, Department of Food Technology, Safety and Health, Faculty of Bioscience Engineering |
| --- | --- |
|  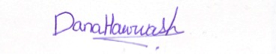 | 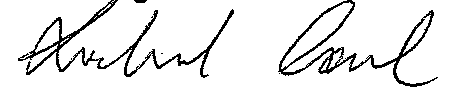 |

1. Lachat, C., et al., *Strengthening the Reporting of Observational Studies in Epidemiology-Nutritional Epidemiology (STROBE-nut): An Extension of the STROBE Statement.* PLoS Med, 2016. **13**(6): p. e1002036.
