## Supplementary material for "Assessment of authors understanding of the STROBE-nut reporting guidelines": Annex 3

**Annex 3 Skype Interview questionnaire**

Proposed questions

- How was your experience with using STROBE nut on your published paper?
- How much time did it take you to fill in the table?
- Was it easy to understand what you have to fill in each column?
- How was it for you to fill in the column “partially included”?
- How was your experience with filling in the reasons for exclusion/partial exclusion?
- Was there any item on the STROBE nut list that was difficult to understand? Can you give an example?
- How can we remove the barriers to make the guidelines more users friendly?

Open-ended

- What is the added value for using STROBE nut for you? (Enriched manuscript, more informative)
- Would you use reporting guidelines e.g STROBE nut on your next manuscript?
- After your experience with STROBE nut application, would you consider the application of other reporting guidelines? If yes, what motivates you to use reporting guidelines?
- Is there anything else you’d like to tell me?
- Can I contact you later in case you have additional questions?
