## Supplementary material for "Assessment of authors understanding of the STROBE-nut reporting guidelines": Annex 4

| Item | Problem | Solution according to participant |
| --- | --- | --- |
| STROBE 1 and nut 1 | “I can understand what these items refer to, but when I was trying to fill in the three choices it was very difficult” |  |
| Nut 5 | “I thought this might not be applicable. What kind of settings? Hospital? For me it was just general population of adults… we didn’t account for cross-countries differences” |  |
| Nut 7.2 | “By nutrition property do you mean the validation or relevance to nutrition, as it wasn’t really clear what you meant by nutritional properties, so when using dietary indices describe the methods used to obtain them, and the nutritional properties, I guess when you are using an index what does that really mean, it was not clear for me. The word property to me means the nutritional property of the food, or what it is trying to convey in terms of its nutritional. I understand what you are trying to say but I do not think that nutritional propriety is the right term, what are they validated on their relevance to nutrition status or dietary quality, because an index in itself doesn’t have a nutrition property, a food does have a nutritional property” | **“I think it is just a language issue,** or maybe you should use nutritional significance or something like that, or what they measure or communicate in terms of nutritional property something like that.” |
| Nut 7.2 | “When you talk about dietary patterns, I wasn’t sure what dietary pattern meant … I didn’t understand dietary pattern meant indicators then I would immediately know.” |  |
| Nut 8.3 | “What do you mean with non-dietary data? What I didn’t understand well here is: is it compulsory to have these non-dietary data? But I think you can have a study where you do not need, or you do not have to…” | “It would have been easier if it was at the end, if applicable” |
| Nut 8.4 | “No but the issue of biomarkers, I mean yea it is not difficult to understand but in terms of my paper, my published paper, we do not have such kind of measurement” |  |
| Nut 8.5 | “Timing of the measurement is not fully clear, you mean at what time it was measured, what kind of example would you say it is important? Because my data was cross sectional so maybe it is more for longitudinal data?” | “An example should be placed in, for example when weight was measured” |
| Nut 8.6 | “Give example of external and internal validation (done by experts outside the group, validated before like the software we have used or quality control inside the group on your own data)” | “Give an example because usually we think about external validation because internal we don’t think much about so it is good to give an example” |
| Nut 11 | “I do not know if this apply to my paper  I wasn’t sure what it meant because I haven’t heard before about N tiles…. I had consumers non consumers and I am not one hundred percent sure it applies to my paper since my categorization is direct beneficiaries and non direct beneficiaries , and not sure if it counts.” |  |
| Nut 12.1 | “The word combine was not clear, because you combine something with something else and here it was not clear what to combine, so I wrote something in the column I thought it is fully reported but honestly I do not know if I understood it correctly.” | “Give an example to make sure we understand…what I wrote in that sense is wrong because I said fully reported on page 3 and 4 but it is not the case” |
| Nut 12.2 | “Could just say it is inapplicable?..i only used diet as outcome and not as exposure…maybe you can up do the calibration or exclude participants for energy intake, I didn’t evaluate the data quantitvely so I cannot report” |  |
| Nut 12.3 | “Are you referring to nutrition data or any data?” | “it is very similar to nut 8.6, I was confused and I was thinking it was only for other measurement than dietary or nutritional measurement.” |
| Nut 20 | “on the relevance of the finding to nutrition. Difficult to grasp. The nutritional relevance of the findings, what do you mean with this, is it relevant on the type of nutrient you report on or the food groups you report on. Relevance van be seen in many dimensions, it could be like okay you have a study on the importance of vitamin A and birth weight which is very relevant. If you increase vitamin A intake, you can have important health outcomes. That might be very relevant in itself, it can be health relevant, is it nutrition relevant yea it is vitamin A so does that makes it less relevant than the fact you find it in a population in high percentage, that is also relevant because it indicates a problem, because how you define relevance. How you define relevance is crucial I mean every research is done because it is relevant. Because irrelevant research should not be published right?” | “Tune it down or add an example” |
| Nut 22.1 and Nut 22.2 | “Item on the availability of data is not specific for only nutritionist, it is more like general. The goal of having this item is different it encourages open science and good research practices and yet it is not specific to nutrition. It fits better in the general strobe than specific for strobe nut because it is for everyone. Same think for 20.1 describe procedures for consent.” | “It is good but it could be required as general strobe item.” |
